# Observational Study on Clinical Features, Treatment and Outcome of COVID 19 in a tertiary care Centre in India- a retrospective case series

**DOI:** 10.1101/2020.08.12.20170282

**Authors:** Raja Bhattacharya, Rohini Ghosh, Manish Kulshrestha, Sampurna Chowdhury, Rishav Mukherjee, Indranil Ray

## Abstract

**Objective:** This study will attempt to explore the demographic profile and outcome in the patients receiving multidisciplinary, personalised approach including use of Broad Spectrum Antivirals - Ivermectin, anti-inflammatory and antioxidants roles of Statins and N-acetyl-cysteine along with Standard of Care (SOC) in hospitalised COVID19 patients in a tertiary care centre.

**Setting:** Inpatient department

**Participants:** 191 COVID-19 patients with laboratory confirmed severe acute respiratory syndrome coronavirus 2 (SARS-CoV-2) infections in the year 2020 between June 14-28, 2020

**Main outcome measures:** The outcome of Interests are:

A. Studying the demographic profile of COVID 19 cases
B. Study the treatment outcomes in terms of death or discharge in patients receiving Ivermectin+N-acetyl-cysteine+Statin along with Standard of care.

**Results:** 148 patients were included in the study. All of them had confirmed COVID19 infection by the rtPCR method. Average age of the patients was 57.57 years (Range = 17 - 88), 49% were male, 51% female. 81% of the patients had at least one or more comorbidities. Most common comorbidities included diabetes (32%), Hypertension (27%),Ischaemic Heart Disease (8%). More comorbidities. The in hospital, Case Fatality Rate was therefore, 1.35 %. The remaining 144 were discharged from the facility after an average 12 days duration of stay.

**Conclusions:** Triple therapy with ivermectin + atorvastatin + N-acetylcysteine can be an useful adjunct to standard of care.

## INTRODUCTION

Since December 2019, the new severe acute respiratory syndrome coronavirus 2 (SARS-CoV-2) which started as an outbreak in Wuhan, China has spread rapidly as a pandemic around the world,^(1)^ causing multi organ dysfunction most prominent of which are coronavirus disease 19 (COVID-19) pneumonia, acute respiratory distress syndrome (ARDS), cardiac injury, liver and renal injury, thrombosis including Pulmonary Embolism and Stroke, and death^(2)^

Although the exact pathophysiology behind COVID19 infection is still unknown, it is primarily known to affect the Respiratory System although other systems including Neurological manifestation are recently coming to light.^(3)^ Structural and functional analysis showed that the spike for SARS-CoV-2 also bound to ACE2 ^(4)(5)(6)^ whose expression was high in lung, heart, ileum, kidney and bladder. In the lung, ACE2 was highly expressed on lung epithelial cells. Patients with severe diseases were reported to have increased plasma concentrations of proinflammatory cytokines, also called a cytokine storm including interleukin (IL)-6, IL-10, granulocyte-colony stimulating factor (G-CSF), monocyte chemoattractant protein 1 (MCP1), macrophage inflammatory protein (MIP)1 *α*, and tumor necrosis factor (TNF)-*α*. ^(7) (8)^ Treatment of SARS-cov-2 is still unknown and mostly symptomatic although clinical application of principles of antiviral therapies are being investigated. The proposed use of repurposed antiviral drugs remains a valid practical consideration. One of the most controversial drugs during the current SARS-CoV-2 pandemic is the well-known oral antimalarial drug hydroxychloroquine (HCQ), routinely used in the treatment of autoimmune diseases like rheumatoid arthritis or lupus. However, all studies that used HCQ with rather contradictory results.

Recently perhaps the most promising treatment strategy has been to use Corticosteroids, in particular Dexamethasone in the RECOVERY trial in reducing mortality. ^(9)^ Other treatment strategies have been to use Low Molecular Heparin to prevent thrombotic complications arising from endothelial dysfunction. ^(10)^

Based on these evidences, Government of India has issued Standard of Care guidelines in managing mild, moderate and severe COVID19 infection which incorporates the use of Oxygenation, restricted fluid therapy, use of anticoagulation either Unfractionated Heparin (UFH) or Low molecular weight heparin (LMWH) and corticosteroid in the form of either Methylprednisolone or Dexamethasone.

This retrospective cohort study aims to describe the demographic profile of hospitalised patients in a tertiary care centre in India and their treatment outcomes when along with Standard of Care, personalised medicine targeting the various pathophysiology of COVID19 was provided. These include the use of antivirals, anti-inflammatory and antioxidants - Ivermectin, Atorvastatin and N-acetylcysteine.

Ivermectin is already known to have broad-spectrum anti-viral activity *in vitro* ^(11-14)^. It inhibits SARS-CoV-2 in vitro, with a single dose to Vero-hSLAM cells 2 h post infection with SARS-CoV-2 and achieves ~5000-fold reduction in viral RNA at 48 h. Ivermectin therefore warrants further investigation for possible benefits in humans. ^(15)^ Based on the promising in vitro data, single dose Ivermectin 12mg was added to standard of care.

As a cure remains elusive, the current management of patients with COVID-19 infection focuses mostly on supportive care with the most severe cases often requiring mechanical ventilation and standard care for ARDS patients of any cause. However, increasingly data is coming to light that ARDS in COVID19 differs from other causes in multiple ways. ^(16)^ ARDS in COVID 19 is very sensitive to PEEP as it has relatively good lung compliance despite poor oxygenation, there is lack of pulmonary vasoconstriction and significant shunting, and thrombotic microangiopathy^(17)(18)(19)^. Some authors suggest that vascular endothelial dysfunction plays a vital role in the pathogenesis of COVID-19 infections ^(17)^. Thus it is postulated that statin treatment may improve endothelial and vascular function in these patients.

In fact, such a combination of statin/ARB treatments has been used previously in some centres during the Ebola outbreak in West Africa ^(20)^. Thus a similar approach has been suggested by Fedson et al ^(21)^ for COVID 19 patients. Statins are known for their pleiotropic anti-inflammatory, antithrombotic and immunomodulatory effects. Based on pathophysiology and understanding of its associated coagulopathy, endothelial dysfunction, and dysregulated inflammation., researchers proposed that statins might mitigate the effects of COVID-19 infection in selected patients. Hence, the decision was taken to add low dose Atorvastatin 10 mg was added to SOC.

Another promising drug due to its antioxidant property was N-acetyl-cysteine. N-acetylcysteine (NAC) is a precursor of glutathione ^(22)^ and acts as a powerful antioxidant (30) and free radical scavenger in the body. There have been several clinical trials investigating the use of NAC in respiratory illness in humans. Intravenous NAC has been used clinically for the treatment of ARDS ^(23)^. In both *in vivo* and human trials, nebulized NAC may improve arterial oxygen tension ^(24), (25)^; and attenuate pulmonary fibrosis ^(26)^, and ARDS ^(27)^.

Thus in this tertiary care hospital adjuvant treatment with Ivermectin + Atorvastatin + NAC was undertaken along with Standard of Care. This study aims to describe the demographic profile and treatment outcomes.

## METHODS

### STUDY DESIGN

This study was done in the inpatient department of designated COVID19 facility of Medical College Kolkata. Hospital records of patients admitted between 14.06.2020 - 21.06.2020 were accessed and daily round data along with severity of disease, medication received and treatment outcome were noted for all patients (n = 148). No sample size calculation was carried out.

This is a retrospective case series aimed at describing patient profile and treatment outcome of patients admitted with COVID 19 infection.

### PATIENT SELECTION, PROCEDURE AND TREATMENT

All patients were rt-PCR confirmed COVID-19 inpatients older than 18 years at diagnosis were included in the analysis as treatment group.

Based on clinical parameters each patient was designated a severity of Mild, Moderate or Severe as per Mohfw guidelines ^(28)^.All patients received Standard of Care based on severity as per MOHFW, India Clinical Guidelines (28) and along with that were given Ivermectin single dose, Atorvastatin 10mg daily and injection N-acetyl-cysteine, irrespective of disease severity.

### DATA COLLECTION

Patient data was collected from written health records in the year 2020. Demographic data, as reported by the patient, and a current medical history of hypertension, diabetes, obesity, cardiovascular disease, heart failure, stroke, asthma, COPD, other lung disease, kidney disease, liver disease, autoimmune disease, history of cancer, thyroid disease were collected.

The following vital signs, if available, were collected and documented: heart rate (beats per minute), breaths per minute (BPM), systolic and diastolic blood pressure (mmHg), body temperature (°C), oxygen saturation measured by pulse oximetry (O2 %), body weight (kg), and/or body mass index (BMI).

### STATISTICAL ANALYSIS

Statistical data was collected and entered into MS-Excel spreadsheets and processed using Stata 12 software for Windows. The demographic data, prevalence of different comorbidities, and case fatality rates were calculated and compared with existing data.

## RESULTS

148 patients were included in the study. All of them had confirmed COVID19 infection by the rtPCR method. Average age of the patients was 57.57 years (Range = 17 - 88), 49% were male, 51% female. 81% of the patients had at least 1 or more comorbidities. Most common comorbidities included diabetes (32%), Hypertension (27%),Ischaemic Heart Disease (8%). Other comorbidities included COPD/BA (3%), Chronic Kidney Disease (6.7%), Chronic Liver Disease (2%) and immunocompromised states including HIV and cancer (6%).

Based on the Clinical severity assessment guidelines ^(28)^ 5% patients had severe disease, 27.5% had moderate disease and 67.5% had mild disease.

None of these patients were lost to follow-up for the defined outcome. Only 2 of the 148 patients treated with Standard of Care + Ivermectin + Atorvastatin + N-acetyl-cysteine, expired. The in hospital, Case Fatality Rate was therefore, 1.35 %, which was well below the national average. ^(29)^ The remaining 144 were discharged from the facility after an average 12 days duration of stay. No notable adverse events related to drugs occurred in any of the subjects during this study.

## DISCUSSION

COVID - 19 till date has no proven effective treatment. Symptomatic therapy along with that pathophysiology based treatment protocols have been adopted by the government. Our observations in this study demonstrate there is a role for further disease modulation based therapy using broad spectrum antiviral - Ivermectin, pleiotropic effects of Atorvastatin and the lung protective and antioxidant of N-acetyl-cysteine. This triple therapy along with standard of care has demonstrated a lower case fatality rate without any notable adverse events and can be considered as adjuncts to successful therapy for COVID 19. However, its observational, non randomised nature prevents a causal conclusion and warrants further investigation through larger Randomised Controlled Trials.

## CONCLUSION

Triple therapy with ivermectin + atorvastatin + N-acetylcysteine can be an useful adjunct to standard of care with no apparent adverse effects and on the contrary decrease mortality to 1.35% well below the national average. ^(29)^

## LIMITATIONS

Retrospective case series study findings that have to be validated in prospective controlled clinical trials. There is limited data on COVID-19 outpatient risk stratification and treatment and more definitive studies should be carried out in future. There was no control group and thus regression models and higher statistical analysis could not be done. This study cannot comment on the efficacy and safety of the used triple therapy related to severely ill hospitalized patients.

## OTHER INFORMATION

### ETHICAL CONSIDERATION

The research protocol was approved by the Institutional Ethics Committee, Medical College, Kolkata and all participants gave written informed consent. Data was redacted and anonymized data was stored in a password protected computer.

## Data Availability

NA

## ACKNOWLEDGMENTS

We acknowledge the dedication, commitment, and sacrifice of the staff, providers, and personnel in our institution through the COVID-19 crisis and the suffering and loss of our patients as well as in their families and our community. We would like to extend our gratitude to the Department of Medicine and Department of Microbiology, Medical College, Kolkata for supporting us.

## CO-AUTHOR CONTACT INFORMATION

Raja Bhattacharya ^1^ (88, College street, Kolkata, India.), Rohini Ghosh ^1^ (88, College street, Kolkata, India.), Manish Kulshrestha ^1^ (88, College street, Kolkata, India.), Sampurna Chowdhury ^1^ (88, College street, Kolkata, India.), Rishav Mukherjee ^1^ (88, College street, Kolkata, India.) and Indranil Ray ^1^* (88, College street, Kolkata, India.).

^1^Medical College Kolkata;

*Corresponding author;

